# Practical challenges to the clinical implementation of saliva for SARS-CoV-2 detection

**DOI:** 10.1101/2020.08.27.20170589

**Authors:** Nancy Matic, Aleksandra Stefanovic, Victor Leung, Tanya Lawson, Gordon Ritchie, Lynne Li, Sylvie Champagne, Marc G. Romney, Christopher F. Lowe

## Abstract

Due to global shortages of flocked nasopharyngeal swabs and appropriate viral transport media during the COVID-19 pandemic, alternate diagnostic specimens for SARS-CoV-2 detection are sought. The accuracy and feasibility of saliva samples collected and transported without specialized collection devices or media were evaluated. Saliva demonstrated good concordance with paired nasopharyngeal swabs for SARS-CoV-2 detection in 67/74 cases (90.5%), though barriers to saliva collection were observed in long-term care residents and outbreak settings. SARS-CoV-2 RNA was stable in human saliva at room temperature for up to 48 hours after initial specimen collection, informing appropriate transport time and conditions.

## Introduction

Diagnostic testing is a cornerstone of the COVID-19 pandemic response strategy[1], yet the establishment of laboratory testing which is accurate, practical, and scalable to meet demand for public health surveillance measures has been a considerable challenge. Nasopharyngeal swabs are the preferred specimen type over throat swabs due to superior sensitivity[2,3], and over nasal aspirates due to lower risk of aerosol generation. Flocked nasopharyngeal swabs are designed to maximize mucosal contact and more efficiently release contents into the testing medium[4,5]; however, due to global demand during the pandemic response, a reliable supply of high-quality, flocked swabs and appropriate viral transport medium has been difficult to procure.

Furthermore, studies describe variable collection quality of nasopharyngeal specimens leading to diminished sensitivity and potential false-negative results for SARS-CoV-2[6-8]. Although samples from the lower respiratory tract such as sputum or bronchoalveolar lavage have been widely used[9-11], only a subset of patients under investigation for COVID-19 are able to expectorate sputum or undergo invasive bronchoscopic procedures.

Saliva has been described as an acceptable alternative diagnostic specimen for detection of common respiratory viruses[12-15], and more recently, SARS-CoV-2[6,16-19]. Salivary gland epithelial cells have demonstrated high expression of ACE2 receptors[20,21], which may enhance the replication of SARS-CoV-2 at this site. Collection techniques vary widely in the published medical literature, and include: passively drooling into a 50mL Falcon tube for one minute[16]; coughing 0.5-1mL of saliva from the back of the throat into a sterile container and adding 2mL of viral transport media upon receipt in the laboratory[17,18]; repeatedly spitting into a sterile container upon waking in the morning prior to eating, drinking, or brushing teeth[6]; or pooling 1-2mL of saliva into a container with addition of liquid Amies media in a 1:1 ratio upon receipt in the laboratory[19].

Saliva collection techniques which require transport media or specialized collection devices are problematic during a pandemic. Distribution of pre-packaged kits may be costly and impractical for community settings, and the reliable supply of necessary materials will continue to be a global challenge. Specialized saliva collection devices typically marketed for biochemical tests[22] may contain components such as cotton which can inhibit nucleic acid amplification. Furthermore, it is uncertain in the current medical literature whether the enzymatic properties of human saliva will readily degrade intact virus particles from infected patients, as opposed to naked viral RNA[23,24]. Commercial RNA stabilization solutions have been marketed to preserve cellular RNA in various specimen types, yet have also been described to decrease efficiency of nucleic acid extraction[25,26]. Clinical diagnostic laboratories require immediate guidance for implementing practical and feasible methods of saliva collection, transport and processing for SARS-CoV-2 detection.

## Materials and Methods

From March-May 2020, saliva and nasopharyngeal swabs were received from patients under investigation for COVID-19 from various clinical settings: symptomatic patients requiring admission to a tertiary acute care hospital; residents of long-term care (LTC) facilities with known COVID-19 outbreaks; healthcare workers; and mildly symptomatic outpatients including household contacts of known positive cases. After the nasopharyngeal swab was collected, patients were asked to provide approximately 1mL of saliva by pooling saliva from their throat and spitting into a sterile screw-top container (Starplex Scientific Inc., Etobicoke, Canada).

Saliva samples were transported to the laboratory at room temperature without addition of transport media, with routine transport times (<24 hours). Upon receipt in the laboratory, samples were diluted 1:2 with sterile phosphate-buffered saline (PBS) and vortexed with glass beads until liquid consistency was achieved. Saliva samples were stored at 4°C until nucleic acid extraction could be performed (<24 hours from time of specimen receipt). Nucleic acid extraction was completed using the MagNA Pure Compact or MagNA Pure 96 System (Roche Molecular Diagnostics, California, USA). Molecular detection of the SARS-CoV-2 envelope (E) gene from saliva samples was performed with the LightMix® ModularDx SARS-CoV (COVID19) E-gene assay (TIB Molbiol, Germany) and LightCycler® Multiplex RNA Virus Master (Roche Molecular Diagnostics, California, USA).

To investigate the optimal transport time of saliva samples without the use of transport media, the stability of SARS-CoV-2 RNA detectable in human saliva samples over time was evaluated. Two patients known to be positive for SARS-CoV-2 and three healthy volunteers provided five saliva samples each. The saliva from volunteers was spiked with 50μL of remnant viral transport media (BD^TM^ universal viral transport system, Maryland, USA) from the nasopharyngeal swabs of SARS-CoV-2 positive patients (Cycle threshold [Ct] value 15-16 for E- gene). Saliva from the patients and volunteers was processed as previously described, but an aliquot of each sample was processed at delayed time points: 0, 12, 24, 36 and 48 hours. In the interim, saliva was stored at room temperature to mirror typical transport conditions. SARS-CoV-2 detection was performed as previously described in triplicate for each sample and mean Ct values were compared over time.

## Results

One hundred and six (106) saliva specimens were received from patients under investigation for COVID-19. Thirty-one saliva specimens (29.2%) had negligible sample volume (<0.5mL). The majority of saliva samples with insufficient volume were from residents and healthcare workers of a LTC facility (30/31, 96.8%). One further specimen was excluded from the analysis due to its appearance as expectorated sputum. On average, the approximate time from saliva collection to the time of laboratory processing was 13 hours for inpatients, 16 hours for outpatients, and 19 hours for samples from LTC facilities. The final analysis included 13/74 samples from inpatients (17.6%), 20/74 samples from LTC residents (27.0%), 28/74 samples from healthcare workers (37.8%), and 13/74 samples from outpatients (17.6%). For the detection of SARS-CoV-2, saliva was concordant with paired nasopharyngeal swabs in 67/74 cases (90.5%); in one case (1.4%), saliva demonstrated detectable SARS-CoV-2 RNA where the paired nasopharyngeal swab was negative.

Stability of SARS-CoV-2 viral RNA in saliva with delayed processing up to 36 hours was demonstrated (Figure 1; mean Ct value for E-gene was 23.75 at time zero versus 24.16 at 36 hours; p-value 0.328, paired *t*-test, GraphPad™). The mean Ct value appeared to demonstrate a significant increase at the last reading near 48 hours (25.76, p-value 0.014, paired *t*-test), although this may be attributed to a single outlier sample (Patient 2), which was collected from an outpatient and had the longest total transport time (53 hours) of all samples.

**Figure 1:**
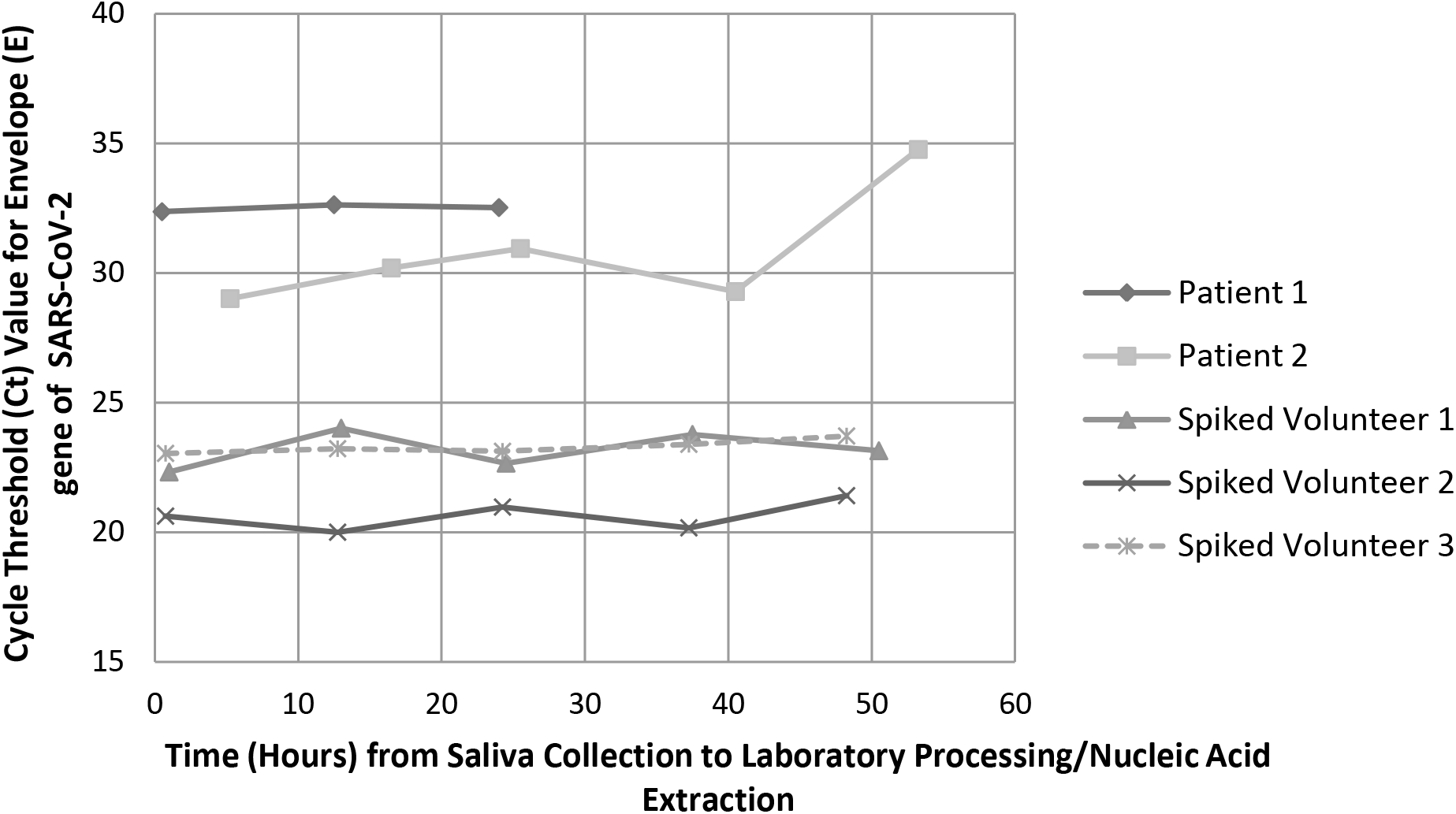
Saliva samples from patients with known COVID-19 infection or from healthy volunteers spiked with viral transport media from the nasopharyngeal swabs of known positive COVID-19 cases were held in the laboratory at room temperature and processed at different time points. The stability of SARS-CoV-2 viral RNA in these samples is represented by the average detected cycle threshold (Ct) value of the Envelope (E) gene of SARS-CoV-2 tested in triplicate and plotted over time. Patient 1 was unable to provide sufficient sample volume for five readings and was only tested up to 24 hours after the initial time of collection.

## Discussion

As a diagnostic specimen for SARS-CoV-2 detection, saliva performed well and demonstrated good general concordance with nasopharyngeal swabs. Although a minority of saliva samples (6/74, 8.1%) were discordantly negative for SARS-CoV-2 RNA, the paired nasopharyngeal swabs in these cases had evidence of low viral loads (late Ct values in the range of 31-39), suggesting the decrease in sensitivity may be attributable to viral loads near the limit of detection of the assay rather than the inherent properties of saliva.

Pre-analytical and analytical factors must be carefully considered before implementation of saliva as a routine diagnostic specimen. Nearly 30% of saliva specimens in this study were of insufficient volume. Although strategies to improve saliva collection have been described[6], LTC residents may face particular challenges due to xerostomia, inability to follow commands, or physical barriers with dentures. Furthermore, saliva may not be suitable for mass testing in outbreak settings where sufficient time or instructions may not be provided. Importantly, the findings from this study indicate saliva without transport media can be in transit at room temperature for up to 48 hours prior to laboratory processing without losing diagnostic yield.

The amount of saliva processing required in the laboratory was significant. Manual labour by laboratory technologists is required to decrease viscosity of saliva and ensure compatibility with laboratory instruments, which has added operational cost. Addition of 1:2 PBS dilutes the specimen and may affect diagnostic sensitivity. However, treatment of saliva in this manner is necessary, as previous experiences in our laboratory revealed nearly all saliva samples demonstrated inhibition with the LightMix® assay when run neat, requiring repeat testing and delaying turn-around-time.

This study had several limitations. Firstly, due to a limited number of samples per each patient group, we are not able to make inferences on performance in any one particular setting. Samples were collected by various healthcare workers without specific training or instruction in saliva collection, and submitted from diverse locations leading to variable transport times. However, these factors do provide a realistic view of the utility of saliva samples in clinical settings. The evaluation of the stability of SARS-CoV-2 viral RNA in human saliva samples without transport media was limited to 48 hours by the available volume of saliva from patients and volunteers. However, the vast majority of samples received by our laboratory for SARS-CoV-2 detection are processed in less than 48 hours, and thus these results have relevance to many clinical diagnostic laboratories.

Further study may include establishment of a reliable method for saliva collection in patient populations with barriers (e.g., LTC residents, intubated patients in critical care units), and optimization of high-throughput automated laboratory instruments to accommodate these highly viscous specimens collected as part of mass surveillance measures.

## Data Availability

This article is a focused, brief report with all relevant data included in the manuscript.

## Acknowledgements

We sincerely extend our gratitude to the patients and volunteers who contributed samples to this study as part of the COVID-19 response. We thank Dr. Michael Schwandt and Noah Reich for their contributions to this project. We are also indebted to our medical laboratory technologists who are highly committed to patient care and laboratory quality improvement.

The authors have no relevant conflicts of interest to declare.

